# Prognostic refinement of NSMP high-risk endometrial cancers using oestrogen receptor immunohistochemistry

**DOI:** 10.1101/2022.09.13.22279853

**Authors:** Lisa Vermij, Jan J. Jobsen, Alicia León-Castillo, Mariel Brinkhuis, Suzan Roothaan, Melanie E. Powell, Stephanie M. de Boer, Pearly Khaw, Linda R. Mileshkin, Anthony Fyles, Alexandra Leary, Catherine Genestie, Ina M. Jürgenliemk-Schulz, Emma J. Crosbie, Helen J. Mackay, Hans. W. Nijman, Remi A. Nout, Vincent T.H.B.M. Smit, Carien L. Creutzberg, Nanda Horeweg, Tjalling Bosse, TransPORTEC consortium

**Affiliations:** Department of Pathology, Leiden University Medical Center, Leiden, the Netherlands; Department of Radiation Oncology, Medisch Spectrum Twente, Enschede, the Netherlands; Department of Pathology, Laboratorium Pathologie Oost-Nederland, Hengelo, the Netherlands; Department of Clinical Oncology, Barts and The London NHS Trust, London, UK; Department of Radiation Oncology, Leiden University Medical Center, Leiden, the Netherlands; Division of Radiation Oncology, Peter MacCallum Cancer Centre, Melbourne, Victoria, Australia; Division of Cancer Medicine, Peter MacCallum Cancer Centre, Melbourne, Victoria, Australia; Department of Radiation Oncology; Princess Margaret Cancer Centre, Toronto, Ontario, Canada; Department of Medical Oncology, Gustave Roussy, Villejuif, France; Department of Pathology, Gustave Roussy, Villejuif, France; Department of Radiation Oncology, University Medical Center Utrecht, Utrecht, the Netherlands; Gynaecological Oncology Research Group, Division of Cancer Sciences, Faculty of Biology, Medicine and Health, University of Manchester, St Mary’s Hospital, Manchester, UK; Department of Obstetrics and Gynaecology, St Mary’s Hospital, Manchester University NHS Foundation Trust, Manchester Academic Health Science Centre, Manchester, UK; Department of Medical Oncology, Odette Cancer Center, Sunnybrook Health Sciences Centre, Toronto, Ontario, Canada; Department of Gynaecology, University Medical Center Groningen, Groningen, the Netherlands

## Abstract

**Background:** Risk-assessment of endometrial cancer (EC) is based on clinicopathological factors and molecular subgroup. It is unclear whether adding hormone receptor expression, L1CAM expression or *CTNNB1* status yields prognostic refinement.

**Methods:** Paraffin-embedded tumour samples of women with high-risk EC (HR-EC) from the PORTEC-3 trial (n=424), and a Dutch prospective clinical cohort called MST (n=256), were used. All cases were molecularly classified. Expression of L1CAM, ER and PR were analysed by whole-slide immunohistochemistry and *CTNNB1* mutations were assessed with a next-generation sequencing. Kaplan-Meier method, log-rank tests and Cox’s proportional hazard models were used for survival analysis.

**Results:** In total, 649 HR-EC were included. No independent prognostic value of ER, PR, L1CAM and *CTNNB1* was found, while age, stage, and adjuvant chemotherapy had an independent impact on risk of recurrence. Subgroup-analysis showed that only in NSMP HR-EC, ER-positivity was independently associated with a reduced risk of recurrence (HR 0.33, 95%CI 0.15-0.75).

**Conclusions:** ER-positivity is a strong favourable prognostic factor in NSMP HR-EC and identifies a homogeneous subgroup of NSMP tumours. ER-positive NSMP EC may be regarded as a novel fifth molecular subgroup. Assessment of ER status in high-risk NSMP EC is feasible in clinical practice and could improve risk stratification and treatment.

## Introduction

Endometrial cancer (EC) is the most common gynaecological malignancy in postmenopausal women.^1^ Although the majority of patients present with early-stage disease and have a good prognosis, 15-20% of women with EC have unfavourable disease characteristics that are associated with an increased risk of distant metastases and EC-related death.^2-4^ In the 2016 ESMO-ESGO-ESTRO guideline, high-risk EC was defined as stage I, grade 3 endometrioid EC (EEC) with deep invasion, stage II or III EEC, or non-endometrioid EC (NEEC).^5^ For these patients, adjuvant pelvic external beam radiotherapy (EBRT) was the standard of care to improve locoregional control.^5^ The randomized PORTEC-3 clinical trial showed that the addition of adjuvant chemotherapy to EBRT (CTRT) increased overall survival (OS) and failure-free survival (FFS) of patients with high-risk EC by 5% and 7% at 5 years, respectively.^6,7^ The greatest OS benefit of CTRT was observed in stage III EC and serous carcinomas (SEC).^7^ Unfortunately, histotype and grade assignment of EC is subject to substantial interobserver variability, hampering the selection of patients that would benefit from CTRT and reducing overtreatment for those who do not.^8^

The EC molecular classification, consisting of the *POLE* ultra-mutated (*POLE*mut), mismatch repair-deficient (MMRd), p53-abnormal (p53abn) and no specific molecular profile (NSMP) molecular subgroups, has repeatedly shown to have strong and independent prognostic value and is also predictive for response to chemotherapy.^9-16^ For this reason, the EC molecular classification was incorporated in the latest European treatment guidelines.^17,18^ The assessment of the molecular classification is encouraged in all EC, especially in high-risk tumours, and a novel risk stratification incorporating the molecular classification has been introduced.^17,18^ All stage I-II *POLE*mut EC are classified as low-risk EC and adjuvant treatment can be safely omitted. In contrast, all p53abn EC with myometrial invasion are now considered high-risk and adjuvant chemotherapy with or without EBRT is recommended.^17,18^ The risk assessment of patients with MMRd and NSMP EC, however, still depends on clinicopathological features such as stage, histotype, FIGO grade and the presence of lymphovascular space invasion (LVSI).

The excellent clinical outcomes of patients with *POLE*mut EC, the intermediate prognosis of MMRd EC and poor survival of p53abn EC has consistently been shown across different cohorts and clinical trials.^9-16^ In contrast, 5-year recurrence-free survival of NSMP EC has varied between intermediate and poor.^9-16^ This heterogeneity in clinical outcomes hampers adequate adjuvant treatment recommendations and suggests biological diversity.

Several molecular alterations that are not included in the current risk stratification have shown to be associated with clinical outcomes in EC, such as *CTNNB1* exon 3 mutations, overexpression of L1CAM, lack of oestrogen receptor (ER) and progesterone receptor (PR) expression, chromosome 1q amplification and other copy number alterations.^10,19-28^ However, the prognostic relevance of these molecular alterations in high-risk EC, in the context of the EC molecular classification, as well as in relation to each other, is less well understood. These molecular alterations may refine the molecular classification and identify subsets of NSMP EC with a distinct prognosis.

Using a large set of molecularly classified high-risk EC from the PORTEC-3 trial and a prospective cohort study, we investigated how ER, PR, L1CAM and *CTNNB1* mutations and established clinicopathologic and molecular risk factors can improve EC risk-assessment.

## Methods

### Patient and tissue selection

This study included patients who participated in the international PORTEC-3 randomized clinical trial, and the prospective clinical cohort of Medisch Spectrum Twente (MST). The design and results of the PORTEC-3 trial have been published previously.^6^ In short, this international phase-III trial randomly assigned 660 eligible patients with high-risk EC (1:1) to postoperative chemoradiotherapy or external beam radiotherapy alone. Inclusion criteria for the trial were: International Federation of Gynecology and Obstetrics (FIGO) 2009 stage IA grade 3 EEC with LVSI; stage IB grade 3 EEC; stage II-IIIC EEC of any grade; or non-endometrioid EC with stages IA (with invasion), IB-IIIC. Upfront central pathology review confirmed the eligibility of all patients.^6^ The presence of LVSI was dichotomously scored as present or absent. The study was approved by the ethics committees at all participating centres. Written informed consent was obtained from all patients.

The prospective cohort study MST included 271 high-risk EC patients who were treated with adjuvant radiotherapy between 1987 and 2015 at Medisch Spectrum Twente, Enschede, The Netherlands. Pathology review was performed by MB, SR and TB to confirm high-risk disease. In contrast to PORTEC-3, LVSI was scored using a 3-tiered scoring system (e.g. no LVSI, focal LVSI, substantial LVSI).^29^ As focal LVSI was not associated with an increased risk of recurrence in previous study^4^, we combined focal LVSI with no LVSI in a final dichotomous LVSI variable. The current study was approved by the Leiden-Den Haag-Delft medical ethics committee, and a waiver for informed consent for the MST cohort was given.

### Immunohistochemistry

Formalin-fixed paraffin-embedded (FFPE) tumour material was available for molecular analyses from 424 (64.2%) PORTEC-3 and 256 (94.5%) MST patients. Whole slide (4 µm) immunohistochemical (IHC) staining for MMR proteins (MLH1, PMS2, MSH2 and MSH6) and p53 on all PORTEC-3 cases was performed and described previously.^12^ Similar IHC staining and scoring for MMR proteins and p53 were performed on cases from MST. When no slides were available for IHC or MMR IHC failed (n = 11), MSI status was determined using the MSI analysis system, version 1.2 (Promega, Madison, WI). In addition, IHC staining for L1CAM, ER and PR was performed on whole slides for all cases. The percentage of positive staining for L1CAM, ER and PR was noted and a 10% cut-off for positivity was used for all three stains, as this cut-off is commonly used for the assessment of L1CAM, ER and PR expression in EC.^10,22-25,30^ A detailed description of all IHC procedures and scoring is available in the Data Supplement.

### Next generation sequencing

Isolation of tumour DNA for targeted next-generation sequencing (NGS) was performed as described previously.^12^ Samples were sequenced using the AmpliSeq Cancer Hotspot Panel version 5 (PORTEC-3) and version 6 (MST) (Thermo Fisher Scientific, Waltham, MA). The presence of pathogenic somatic mutations was evaluated, considering a minimum coverage of 100 reads and variant allele frequency of 10%. A detailed description of DNA isolation and sequencing is available in the Data Supplement. When no slides were available for IHC or p53 IHC failed (n = 20), the final p53 status was determined by the *TP53* mutation status. In cases with failed NGS, KASPar competitive allele-specific polymerase chain reaction (LGC Genomics, Berlin, Germany) assays were used to screen for hotspot mutations in *POLE* (including codons 286, 297, 411, 456 and 459) as previously reported.^12^ Evaluation of IHC and sequencing results was performed blinded to each other and patient outcome.

### Statistical Analysis

The primary endpoint was recurrence-free survival (RFS); calculated from the date of randomization (PORTEC-3) or date of start of adjuvant treatment (MST) to the date of the event of interest, or date of the last follow-up in patients without events. Secondary endpoints were locoregional recurrence-free survival (including vaginal and pelvic recurrences), distant metastasis-free survival (including para-aortic, abdominal and other distant recurrences), and disease-specific survival (DSS). For locoregional, distant and overall recurrence-free survival, event-free patients who died due to other causes than EC were censored.

Differences between groups were tested using the χ^2^ test or Fisher’s exact test for categorical variables, and with the Mann-Whitney U test for ordinal and non-normally distributed continuous variables. Median follow-up time was estimated using the reverse Kaplan-Meier method. Survival analyses were performed according to Kaplan-Meier’s method and groups were compared with the log-rank test. Cox’ proportional hazards models were used to evaluate the prognostic value of (established) clinicopathological and molecular features in the complete study population, as well as in the molecular subgroups separately. Step-wise backward likelihood ratio-based variable selection with stratification for cohort was applied to build multivariable models. The relative importance of variables included in the multivariable models was based on the variable’s proportion of the χ2 statistic. Model validation was performed by analysis of discrimination and indices of optimism determined by means of model fitting to 1000 bootstrap resamples. In addition, internal validation using the leave-one-out method was performed by re-estimating on the two cohorts independently. Comparison of fit between multivariable models was performed by means of Akaike’s information criterion (AIC), model concordance (C-statistic) and likelihood ratio test for comparison of nested models. A two-sided *p*-value <0.05 was considered statistically significant. Statistical analyses were performed with SPSS (Statistical Package of Social Science) version 25 (IBM, Armonk, NY, USA) and R (version 3.6.3., https://r-project.org) using the survival package.

## Results

### Clinicopathologic characteristics

Molecular classification was successfully determined in 411 EC from PORTEC-3 and 237 EC from MST, making a total of 648 molecularly classified high-risk EC eligible for analyses (supplementary figure S1). There were no significant differences in patient and tumour characteristics between included and excluded patients (supplementary table S1), except that the included patients more frequently received EBRT and had a slightly lower 5-year overall survival (71.7% vs 77.0%, p = 0.031) compared to the excluded patients (supplementary table S1).

Characteristics of the included patients from PORTEC-3 and MST are shown in table 1. Although MST had inclusion criteria similar to PORTEC-3, minor differences between the cohorts were observed: patients from MST predominantly received EBRT (n=199, 85.0%), and some had carcinosarcomas (n=24, 10.1%). Median follow-up time of the complete cohort was 7.0 years (95% CI 6.7-7.2).

**Table 1.**
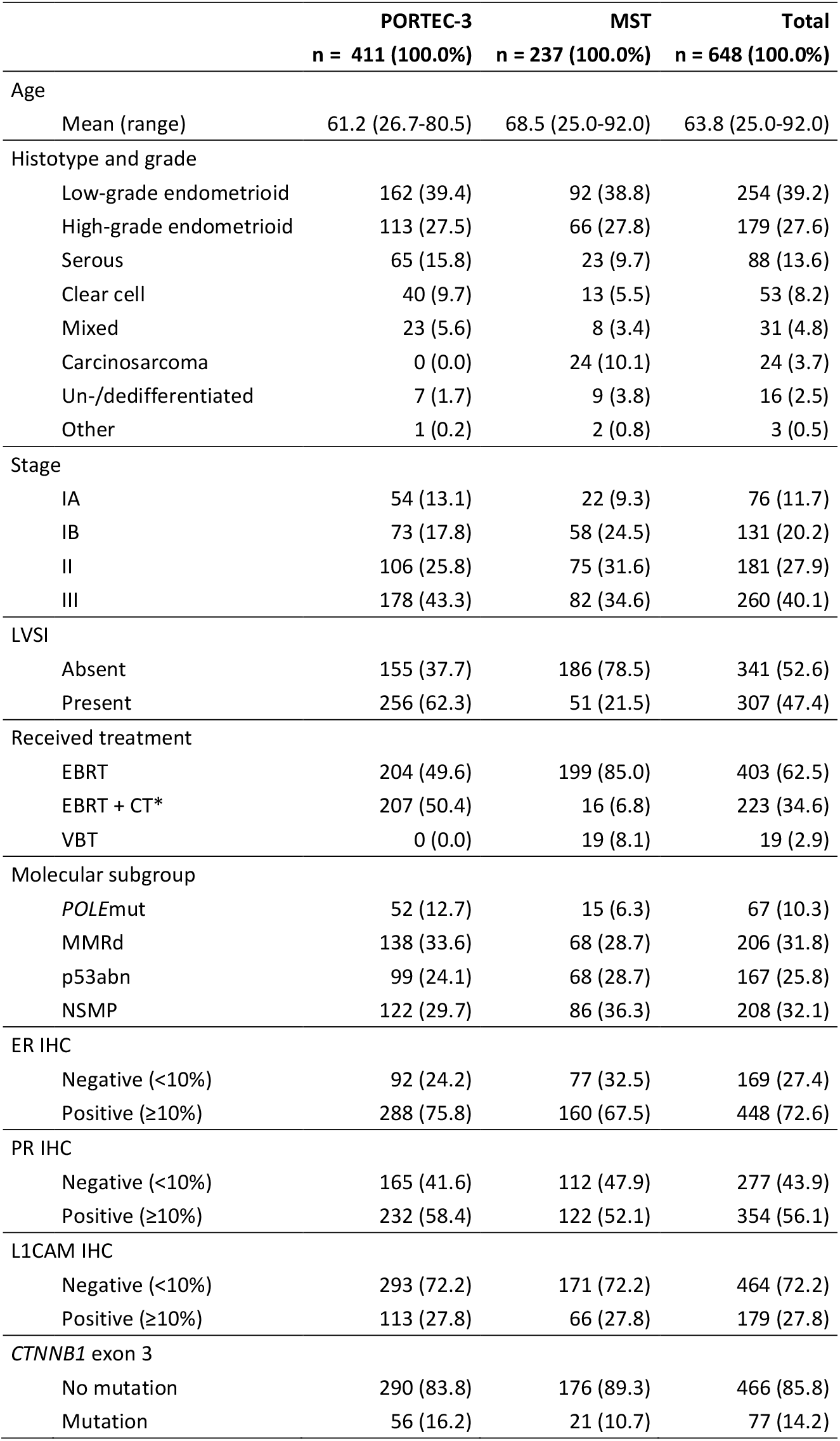
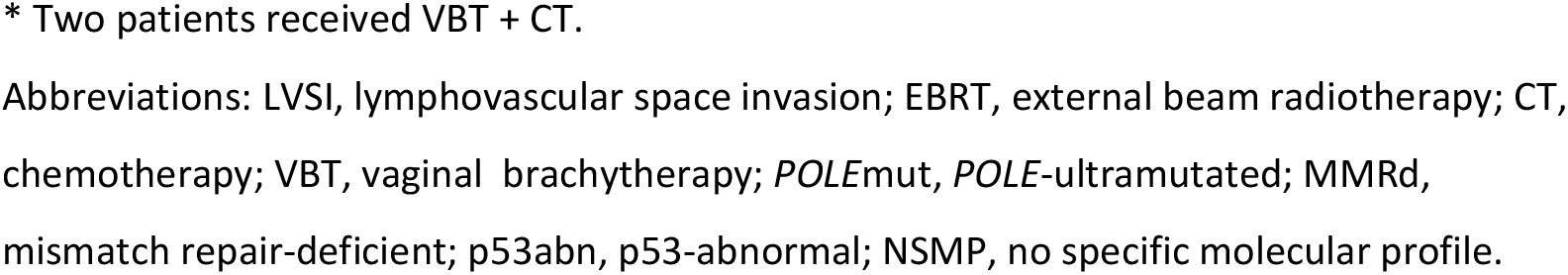
Patient, tumour and treatment characteristics.

|  | **PORTEC-3**<br>**n = 411 (100.0%)** | **MST**<br>**n = 237 (100.0%)** | **Total**<br>**n = 648 (100.0%)** |
| --- | --- | --- | --- |
| Age |  |  |  |
| Mean (range) | 61.2 (26.7-80.5) | 68.5 (25.0-92.0) | 63.8 (25.0-92.0) |
| Histotype and grade |  |  |  |
| Low-grade endometrioid | 162 (39.4) | 92 (38.8) | 254 (39.2) |
| High-grade endometrioid | 113 (27.5) | 66 (27.8) | 179 (27.6) |
| Serous | 65 (15.8) | 23 (9.7) | 88 (13.6) |
| Clear cell | 40 (9.7) | 13 (5.5) | 53 (8.2) |
| Mixed | 23 (5.6) | 8 (3.4) | 31 (4.8) |
| Carcinosarcoma | 0 (0.0) | 24 (10.1) | 24 (3.7) |
| Un-/dedifferentiated | 7 (1.7) | 9 (3.8) | 16 (2.5) |
| Other | 1 (0.2) | 2 (0.8) | 3 (0.5) |
| Stage |  |  |  |
| IA | 54 (13.1) | 22 (9.3) | 76 (11.7) |
| IB | 73 (17.8) | 58 (24.5) | 131 (20.2) |
| II | 106 (25.8) | 75 (31.6) | 181 (27.9) |
| III | 178 (43.3) | 82 (34.6) | 260 (40.1) |
| LVSI |  |  |  |
| Absent | 155 (37.7) | 186 (78.5) | 341 (52.6) |
| Present | 256 (62.3) | 51 (21.5) | 307 (47.4) |
| Received treatment |  |  |  |
| EBRT | 204 (49.6) | 199 (85.0) | 403 (62.5) |
| EBRT + CT* | 207 (50.4) | 16 (6.8) | 223 (34.6) |
| VBT | 0 (0.0) | 19 (8.1) | 19 (2.9) |
| Molecular subgroup |  |  |  |
| *POLE*mut | 52 (12.7) | 15 (6.3) | 67 (10.3) |
| MMRd | 138 (33.6) | 68 (28.7) | 206 (31.8) |
| p53abn | 99 (24.1) | 68 (28.7) | 167 (25.8) |
| NSMP | 122 (29.7) | 86 (36.3) | 208 (32.1) |
| ER IHC |  |  |  |
| Negative (<10%) | 92 (24.2) | 77 (32.5) | 169 (27.4) |
| Positive (≥10%) | 288 (75.8) | 160 (67.5) | 448 (72.6) |
| PR IHC |  |  |  |
| Negative (<10%) | 165 (41.6) | 112 (47.9) | 277 (43.9) |
| Positive (≥10%) | 232 (58.4) | 122 (52.1) | 354 (56.1) |
| L1CAM IHC |  |  |  |
| Negative (<10%) | 293 (72.2) | 171 (72.2) | 464 (72.2) |
| Positive (≥10%) | 113 (27.8) | 66 (27.8) | 179 (27.8) |
| *CTNNB1* exon 3 |  |  |  |
| No mutation | 290 (83.8) | 176 (89.3) | 466 (85.8) |
| Mutation | 56 (16.2) | 21 (10.7) | 77 (14.2) |

\* Two patients received VBT + CT.
Abbreviations: LVSI, lymphovascular space invasion; EBRT, external beam radiotherapy; CT, chemotherapy; VBT, vaginal brachytherapy; *POLE*mut, *POLE*-ultramutated; MMRd, mismatch repair-deficient; p53abn, p53-abnormal; NSMP, no specific molecular profile.

### Molecular and other prognostic factors and correlation with clinical outcome

Prognostic value of the molecular classification for locoregional, distant and overall RFS and CSS was evaluated (figure 1). For all four outcomes, *POLE*mut EC showed an excellent prognosis; even among the 17 patients with stage III *POLE*mut disease, only 1 recurrence was observed. p53abn EC showed the poorest clinical outcomes, while MMRd and NSMP EC had intermediate clinical outcomes. Kaplan-Meier analysis of RFS stratified by cohort is provided in supplementary figure S2.

**Figure 1.**
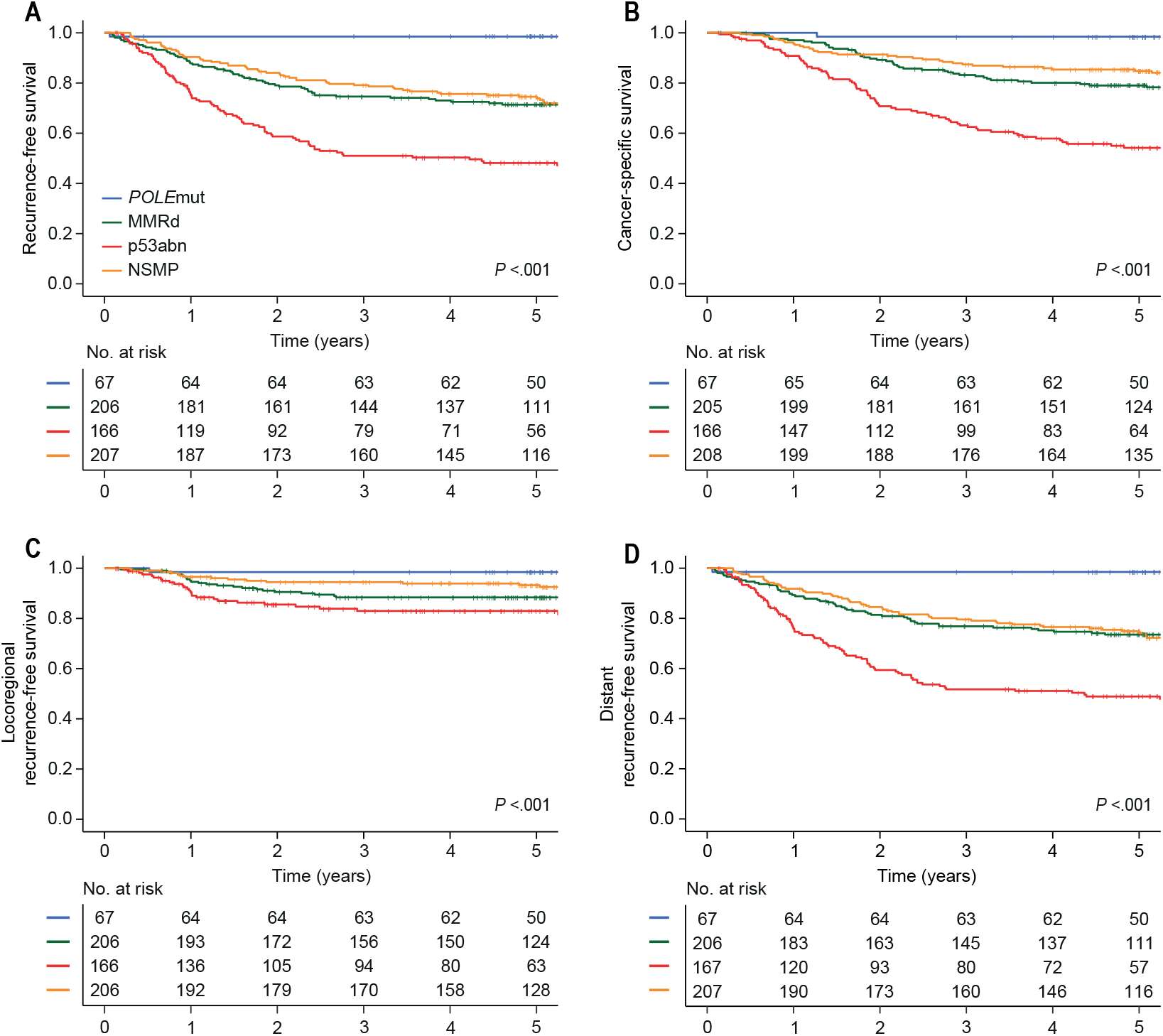
Locoregional, distant and overall recurrence-free survival, and cancer-specific survival for patients with high-risk endometrial cancer (n = 647). Kaplan-Meier survival curves of patients with high-risk endometrial cancer (EC) for (A) locoregional recurrence-free survival (RFS) for patients with *POLE*mut EC (5-year RFS 98.5%), MMRd EC (5-year RFS 88.4%), p53abn EC (5-year RFS 83.0%) and NSMP EC (5-year RFS 93.3%), (B) distant RFS for patients with *POLE*mut EC (5-year RFS 98.5%), MMRd EC (5-year RFS 73.5%), p53abn EC (5-year RFS 48.8%) and NSMP EC (5-year RFS 74.9%), (C) overall RFS for patients with *POLE*mut EC (5-year RFS 98.5%), MMRd EC (5-year RFS 71.4%), p53abn EC (5-year RFS 48.1%) and NSMP EC (5-year RFS 74.5%), and (D) cancer-specific survival for patients with *POLE*mut EC (5-year RFS 98.5%), MMRd EC (5-year RFS 79.1%), p53abn EC (5-year RFS 54.2%), NSMP EC (5-year RFS 84.8%). Abbreviations: *POLE*mut, *POLE*-ultramutated; MMRd, mismatch repair deficient; p53abn, p53-abnormal; NSMP, no specific molecular profile.

Next, we evaluated the prognostic value of ER, PR, L1CAM and *CTNNB1* and established risk factors across all cases (table 2). Independent predictors for lower RFS in multivariable analysis were age at diagnosis above 60 years (HR 1.43, 95% CI 1.02-2.01), stage II (HR 1.78, 95% CI 1.15-2.75) and III disease (HR 3.47, 95% CI 2.37-5.07), and the p53abn molecular subgroup (HR 2.43, 95% CI 1.65-3.57). Adjuvant CTRT and *POLE*mut molecular subgroup were independent predictors for better RFS (HR 0.65, 95% CI 0.47-0.91 and HR 0.11, 95% CI 0.03-0.46, respectively). ER, PR, L1CAM and *CTNNB1* were not found to be predictive of recurrence in multivariable analysis, after correction for clinicopathological risk factors and molecular subgroup.

**Table 2.**
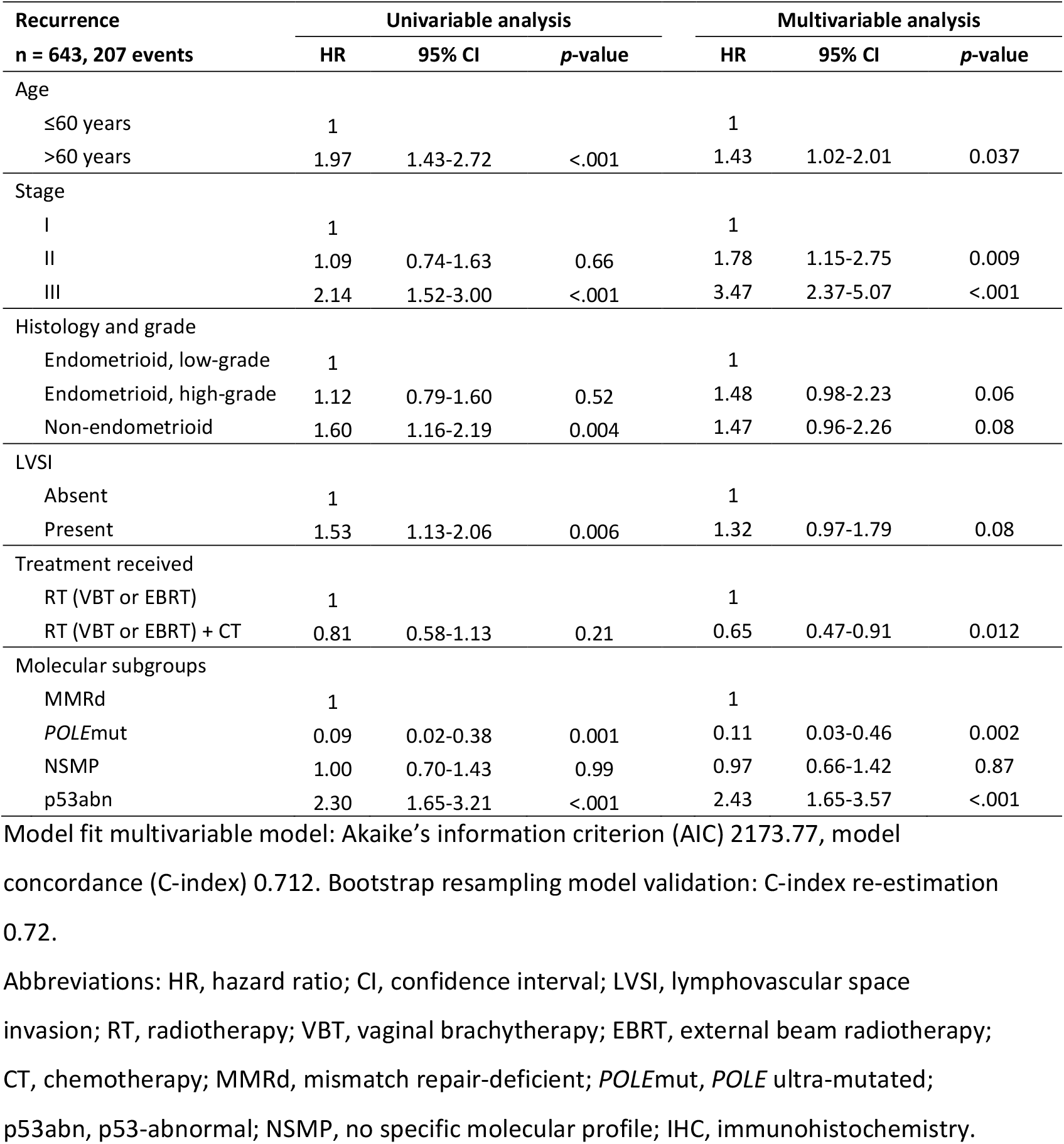
Univariable and multivariable analysis of clinicopathological and molecular features in high-risk endometrial cancer patients.

| Recurrence<br>n = 643, 207 events | Univariable analysis |  |  | Multivariable analysis |  |  |
| --- | --- | --- | --- | --- | --- | --- |
|  | HR | 95% CI | p-value | HR | 95% CI | p-value |
| Age |  |  |  |  |  |  |
| ≤60 years | 1 |  |  | 1 |  |  |
| >60 years | 1.97 | 1.43-2.72 | <.001 | 1.43 | 1.02-2.01 | 0.037 |
| Stage |  |  |  |  |  |  |
| I | 1 |  |  | 1 |  |  |
| II | 1.09 | 0.74-1.63 | 0.66 | 1.78 | 1.15-2.75 | 0.009 |
| III | 2.14 | 1.52-3.00 | <.001 | 3.47 | 2.37-5.07 | <.001 |
| Histology and grade |  |  |  |  |  |  |
| Endometrioid, low-grade | 1 |  |  | 1 |  |  |
| Endometrioid, high-grade | 1.12 | 0.79-1.60 | 0.52 | 1.48 | 0.98-2.23 | 0.06 |
| Non-endometrioid | 1.60 | 1.16-2.19 | 0.004 | 1.47 | 0.96-2.26 | 0.08 |
| LVSI |  |  |  |  |  |  |
| Absent | 1 |  |  | 1 |  |  |
| Present | 1.53 | 1.13-2.06 | 0.006 | 1.32 | 0.97-1.79 | 0.08 |
| Treatment received |  |  |  |  |  |  |
| RT (VBT or EBRT) | 1 |  |  | 1 |  |  |
| RT (VBT or EBRT) + CT | 0.81 | 0.58-1.13 | 0.21 | 0.65 | 0.47-0.91 | 0.012 |
| Molecular subgroups |  |  |  |  |  |  |
| MMRd | 1 |  |  | 1 |  |  |
| *POLE*mut | 0.09 | 0.02-0.38 | 0.001 | 0.11 | 0.03-0.46 | 0.002 |
| NSMP | 1.00 | 0.70-1.43 | 0.99 | 0.97 | 0.66-1.42 | 0.87 |
| p53abn | 2.30 | 1.65-3.21 | <.001 | 2.43 | 1.65-3.57 | <.001 |

Model fit multivariable model: Akaike's information criterion (AIC) 2173.77, model concordance (C-index) 0.712. Bootstrap resampling model validation: C-index re-estimation 0.72.
Abbreviations: HR, hazard ratio; CI, confidence interval; LVSI, lymphovascular space invasion; RT, radiotherapy; VBT, vaginal brachytherapy; EBRT, external beam radiotherapy; CT, chemotherapy; MMRd, mismatch repair-deficient; *POLE*mut, *POLE* ultra-mutated; p53abn, p53-abnormal; NSMP, no specific molecular profile; IHC, immunohistochemistry.

Next, we investigated molecular subgroup-specific prognostic factors (table 3, supplementary table S2). As only 1 patient with a *POLE*mut EC experienced a recurrence, no multivariable analysis was performed for this molecular subgroup.

**Table 3.**
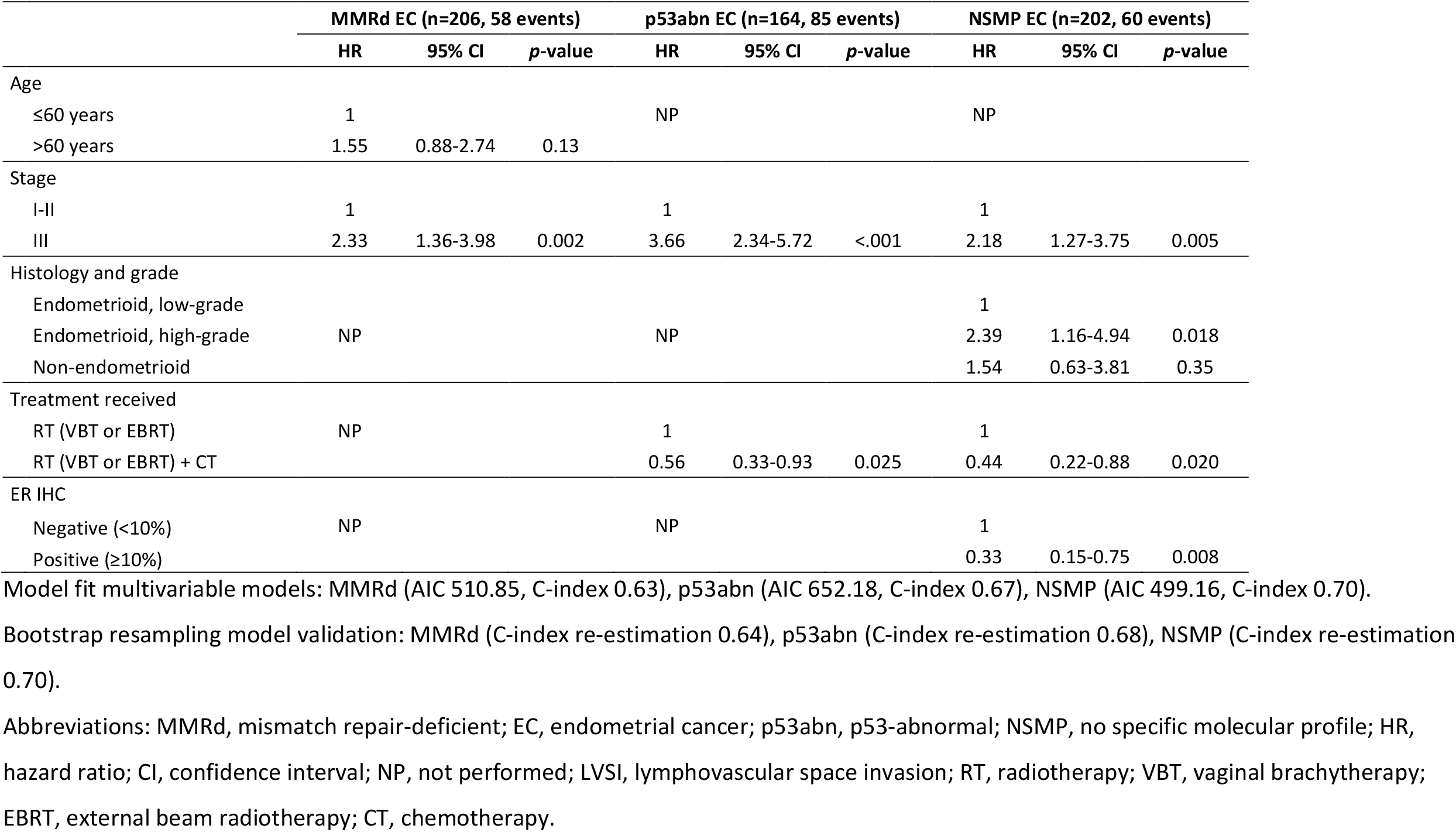
Multivariable analysis of recurrence-free survival including clinicopathological and molecular features for MMRd, p53abn and NSMP endometrial cancers.

|  | MMRd EC (n=206, 58 events) |  |  | p53abn EC (n=164, 85 events) |  |  | NSMP EC (n=202, 60 events) |  |  |
| --- | --- | --- | --- | --- | --- | --- | --- | --- | --- |
|  | HR | 95% CI | p-value | HR | 95% CI | p-value | HR | 95% CI | p-value |
| Age |  |  |  |  |  |  |  |  |  |
| ≤60 years | 1 |  |  | NP |  |  | NP |  |  |
| >60 years | 1.55 | 0.88-2.74 | 0.13 |  |  |  |  |  |  |
| Stage |  |  |  |  |  |  |  |  |  |
| I-II | 1 |  |  | 1 |  |  | 1 |  |  |
| III | 2.33 | 1.36-3.98 | 0.002 | 3.66 | 2.34-5.72 | <.001 | 2.18 | 1.27-3.75 | 0.005 |
| Histology and grade |  |  |  |  |  |  |  |  |  |
| Endometrioid, low-grade |  |  |  |  |  |  | 1 |  |  |
| Endometrioid, high-grade | NP |  |  | NP |  |  | 2.39 | 1.16-4.94 | 0.018 |
| Non-endometrioid |  |  |  |  |  |  | 1.54 | 0.63-3.81 | 0.35 |
| Treatment received |  |  |  |  |  |  |  |  |  |
| RT (VBT or EBRT) | NP |  |  | 1 |  |  | 1 |  |  |
| RT (VBT or EBRT) + CT |  |  |  | 0.56 | 0.33-0.93 | 0.025 | 0.44 | 0.22-0.88 | 0.020 |
| ER IHC |  |  |  |  |  |  |  |  |  |
| Negative (<10%) | NP |  |  | NP |  |  | 1 |  |  |
| Positive (≥10%) |  |  |  |  |  |  | 0.33 | 0.15-0.75 | 0.008 |

Model fit multivariable models: MMRd (AIC 510.85, C-index 0.63), p53abn (AIC 652.18, C-index 0.67), NSMP (AIC 499.16, C-index 0.70).
Bootstrap resampling model validation: MMRd (C-index re-estimation 0.64), p53abn (C-index re-estimation 0.68), NSMP (C-index re-estimation 0.70).
Abbreviations: MMRd, mismatch repair-deficient; EC, endometrial cancer; p53abn, p53-abnormal; NSMP, no specific molecular profile; HR, hazard ratio; CI, confidence interval; NP, not performed; LVSI, lymphovascular space invasion; RT, radiotherapy; VBT, vaginal brachytherapy; EBRT, external beam radiotherapy; CT, chemotherapy.

Among MMRd EC, both uni- and multivariable analyses showed that stage was a significant predictor for recurrence (stage I-II vs III, HR 2.33, 95%CI 1.36-3.98, p=0.002) (table 3, supplementary table S2). Histotype and grade did not have prognostic value within MMRd, as shown in supplementary figure S2. ER, PR, L1CAM and *CTNNB1* were also not associated with recurrence in multivariable analysis of MMRd EC.

Within the subgroup of p53abn EC, uni- and multivariable analyses showed that more advanced stage was significantly associated with recurrence (stage I-II vs III, HR 3.66, 95% CI 2.34-5.72, p<.001) (table 3, supplementary table S2). Furthermore, CTRT was associated with a decreased risk of recurrence compared to RT alone (HR 0.56, 95% CI 0.33-0.93, p=0.025). No prognostic impact of histotype and grade, and ER, PR, L1CAM and *CTNNB1* was found.

Within the subgroup of NSMP EC, ER- and PR-positivity were found to be independently associated with a more favourable RFS (table 3, supplementary table S2). Because ER and PR expression were significantly correlated (Spearman’s rho 0.67, p <.001), we investigated by Kaplan-Meier analysis of RFS whether a combination of ER and PR status was relevant for prognosis. Figure 2 shows that women with ER-positive NSMP EC have a better RFS than those with ER-negative NSMP EC, regardless of the PR status. Of note, no ER-negative and PR-positive NSMP EC were encountered. Further exploration of the relation of ER, PR and the landscape of pathological and molecular features of NSMP EC revealed that ER negativity, rather than PR negativity, was associated with aggressive characteristics such as high-grade, non-endometrioid histology and L1CAM overexpression (figure 3). Based on these findings, ER and not PR status was analysed by multivariable regression, which showed strong prognostic impact on RFS, corrected for stage, tumour grade and adjuvant therapy (table 3). Internal validation confirmed the prognostic effect of ER in NSMP EC (supplementary table S3). To evaluate the chosen cut-off of 10% for ER positivity within NSMP EC we performed a Kaplan-Meier analysis for RFS by percentage of ER expression in tumour tissue, which showed that a threshold of 10% has more discriminative power than a threshold of 1% (supplementary figure S4).

**Figure 2.**
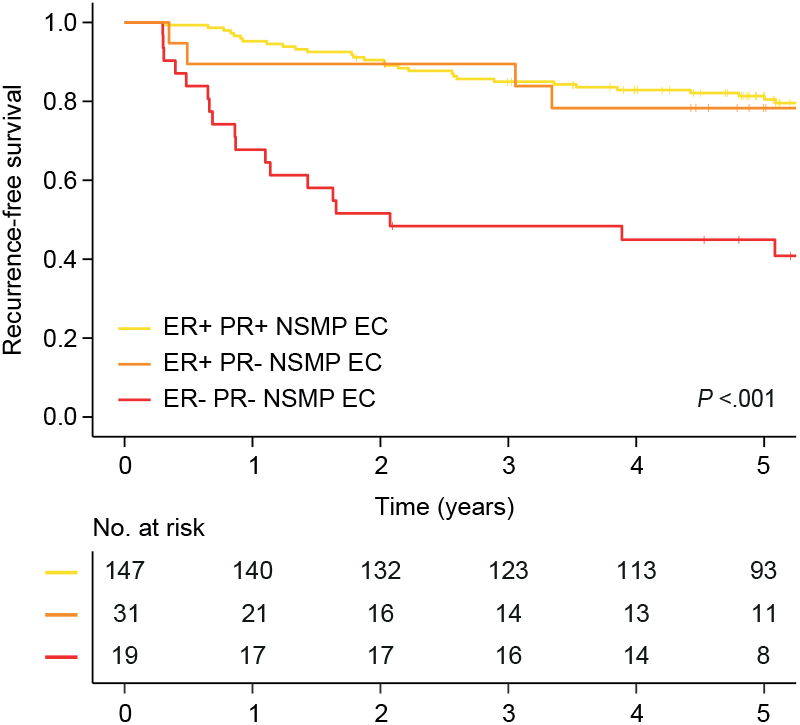
Recurrence-free survival for patients with NSMP high-risk endometrial cancer by ER and PR expression. Kaplan-Meier survival curves of patients with NSMP high-risk endometrial cancer for recurrence-free survival by ER and PR expression.

**Figure 3.**
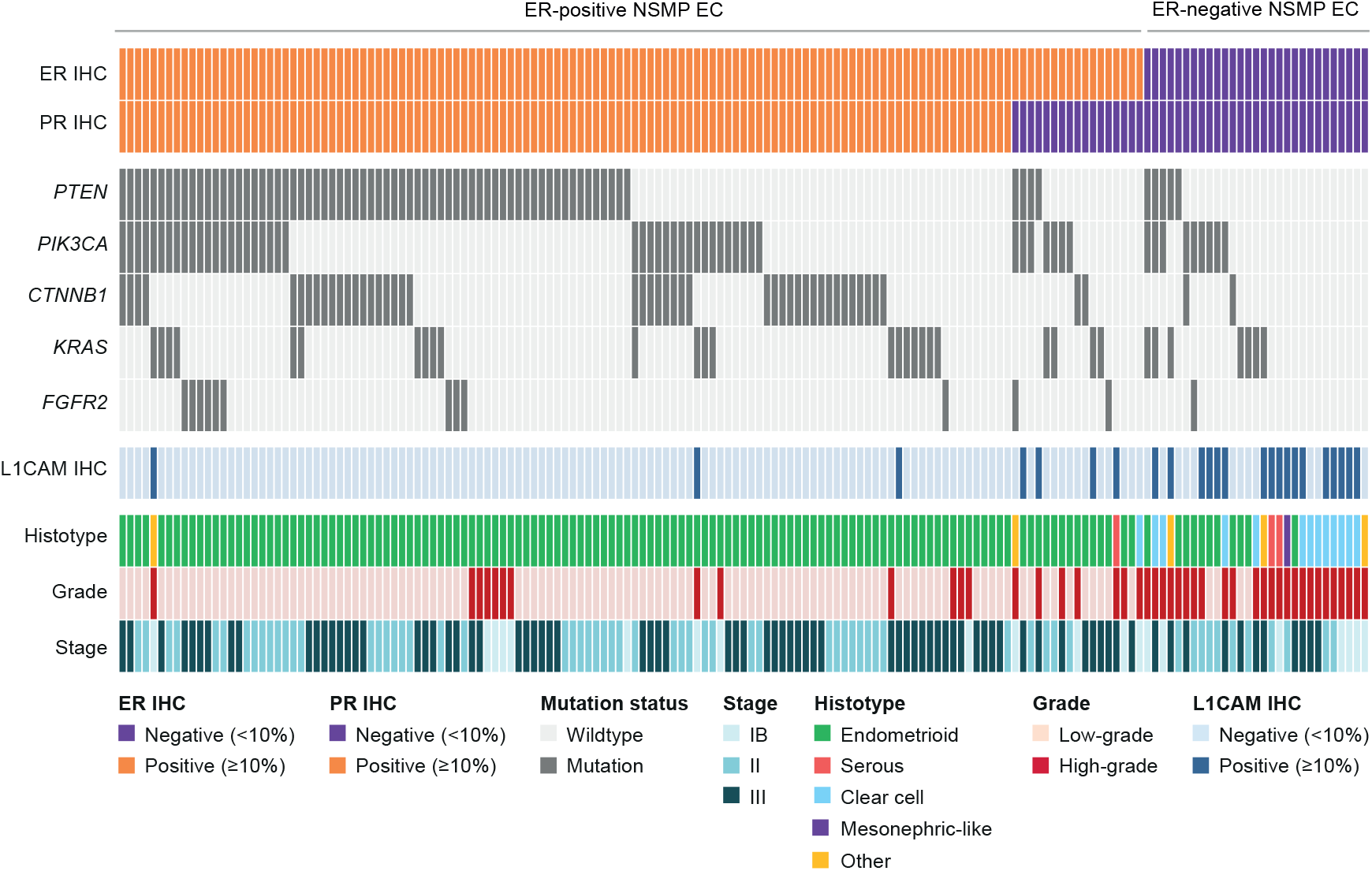
Histopathological and molecular characteristics of NSMP high-risk endometrial cancers. Histopathological and molecular landscape depicting ER and PR status, the most frequently mutated genes, histotype and grade assignment and L1CAM status of NSMP high-risk endometrial cancers (n = 161) with successful ER, PR and L1CAM immunohistochemistry and next generation sequencing. Abbreviations: IHC, immunohistochemistry.

Finally, we evaluated differences in adjuvant treatment effect (CTRT vs. RT) between ER-positive and ER-negative NSMP EC (supplementary figure S5). Both patients with ER-positive and ER-negative NSMP EC appeared to have a small non-significant benefit of CTRT compared to RT alone.

### Prognostic refinement of the EC molecular classification

We tested the incorporation of ER-negative NSMP and ER-positive NSMP tumours as separate molecular subgroups by comparing our multivariable model for RFS, including the molecular classifier with four subgroups (Table 2), with the same model including a five-class molecular classifier (dividing NSMP into ER-positive and ER-negative; supplementary table S4). This improved model fit (AIC 2173.77 vs. 2162.38, C-index 0.712 vs. 0.726, p<.001). In the multivariable model with five molecular subgroups, the ER-negative NSMP group was independently associated with a significantly worse RFS (HR 2.27, 95% CI 1.33-3.90, p=0.003), while the NSMP ER-positive group was not (HR 0.69 95% CI 0.45-1.06, p=0.09), compared to the reference group MMRd.

## Discussion

In this comprehensive analysis of 648 high-risk EC, we evaluated the prognostic value of ER, PR, L1CAM and *CTNNB1* mutations and established clinicopathologic and molecular risk factors in one of the largest cohorts of molecularly classified high-risk endometrial cancers worldwide. Overall, no independent prognostic value of ER, PR, L1CAM and *CTNNB1* was found, while the known independent impact of age, stage, the EC molecular classification and CTRT on risk of recurrence was confirmed. Within the NSMP molecular subgroup prognosis was clearly different by stage and grade, and women with ER-positive tumours had a substantially reduced risk of recurrence compared to those with ER-negative tumours. ER status, which can easily be assessed in routine diagnostics with immunohistochemistry, has the potential to refine risk stratification of women with high-risk NSMP EC.

In our complete study cohort we did not find independent prognostic relevance of ER, PR, L1CAM and *CTNNB1* status. Subgroup-analysis by molecular subgroup did not show prognostic relevance of PR, L1CAM and *CTNNB1* status either. Importantly, ER status was an important predictor for RFS specifically in NSMP EC, but not in *POLE*mut, MMRd and p53abn EC. ER positivity appeared to identify a largely homogeneous group of NSMP EC with (low-grade) endometrioid histology, frequent alterations in the PI3K- and Wnt-signalling pathways and relatively favourable clinical outcomes. In contrast, the small group of ER-negative NSMP EC remained morphologically and molecularly heterogeneous, albeit all associated with more aggressive features such as non-endometrioid histology and poor clinical outcomes. Internal validation confirmed the prognostic effect of ER in NSMP EC. Given these findings, we propose to include ER into the WHO diagnostic algorithm for the molecular classification resulting in a novel molecular subgroup (ER-positive NSMP EC) (supplementary figure S6). This proposed molecular classification with 5 subgroups significantly improved prognostication in our cohorts of high-risk EC, with a clinically relevant difference in 5-year RFS between ER-positive and ER-negative NSMP EC (80.9% vs. 45.3% respectively, p<.001).

The small group of ER-negative NSMP EC remains morphologically and molecularly heterogeneous, albeit with a common association of more aggressive features. A notable proportion of ER-negative NSMP tumours in our cohort were clear cell carcinomas. This rare type of endometrial cancer is generally associated with aggressive clinical behaviour, although recent studies suggest that this is molecular subgroup-dependent, with only NSMP and p53abn clear cell carcinomas having poor clinical outcomes.^31,32^ Currently, NSMP clear cell carcinomas are excluded from the prognostic risk groups of the European clinical guidelines due to insufficient evidence.^17,33^ Incorporating ER status of NSMP EC into the prognostic risk groups will decrease the number of patients that cannot be classified. Mesonephric-like carcinoma is another rare and aggressive type of EC that has only recently been recognized. These tumours are often morphologically mistaken for more common EC histotypes, such as low-grade endometrioid EC. Mesonephric-like carcinomas show intact MMR proteins and wildtype p53 expression and are thus frequently molecularly classified as NSMP EC. They are typically characterised by *KRAS* mutations, absence of *PTEN* gene alterations, chromosome 1q gains, expression of TTF-1 and/or GATA-3, and lack of ER expression.^34-37^ Correct identification of mesonephric-like carcinomas is crucial because of their poor clinical outcomes, including frequent metastases to the lungs, especially when compared to low-grade endometrioid EC.^38^ Finally, some ER-negative NSMP tumours may have high levels of copy number alterations without p53 abnormalities. In the TCGA analyses, pathogenic *TP53* mutations were present in 90% of copy number-high tumours.^39^ As p53 IHC and/or *TP53* mutation analysis are used as surrogate markers for the identification of copy number-high tumours, a small proportion will be classified as NSMP EC. Previous studies showed that relatively high copy number alterations, including chromosome 1q gain/amplification, is associated with negative ER expression and adverse clinical outcomes in NSMP EC.^27,28^

There is currently no consensus about the IHC expression threshold to define ER positivity in EC. We used a 10% cut-off, as this is a commonly used threshold in EC.^23-25,30^ However, some studies use a 1% threshold ^40^ which is also used for selecting patients for hormonal therapy in advanced EC.^17^ Our analysis of a large cohort of high-risk NSMP EC showed that using a 10% threshold yields the best distinction in terms of prognosis. Future studies are warranted to validate this 10% cut-off for prediction of prognosis and response to hormonal therapy in EC patients.

The recent incorporation of the EC molecular classification into the clinical guidelines has improved the risk stratification of EC patients.^17,18^ For NSMP EC patients, risk group assignment depends on stage, histotype, grade and LVSI status. Our results suggest that the addition of ER status can improve risk stratification of patients with NSMP EC. ER-negative NSMP tumours showed poor clinical outcomes, even comparable to p53abn EC, independent of other risk factors. Another study, including only high-grade endometrioid and non-endometrioid EC, reported similar poor clinical outcomes for NSMP EC.^13^ In this study, half of the NSMP EC were non-endometrioid (16% serous EC and 33% clear cell carcinomas) and plausibly ER-negative. It is, therefore, likely that all ER-negative NSMP EC have a high risk of recurrence. Our proposed prognostic stratification of NSMP into ER-positive and ER-negative NSMP EC should be evaluated in future studies that also include lower risk NSMP EC.

In addition to ER status, the tumour stage, histotype and grade were independent predictors for recurrence in NSMP EC and may therefore still be relevant in the risk stratification of ER-positive NSMP EC. In *POLE*mut, MMRd, p53abn endometrioid EC tumour grading was not informative. Confirmation of this finding in other cohorts may lead to a simplification in diagnosing and classifying patients in risk groups by limiting tumour grading to NSMP EEC. Remarkably, we found no significant independent prognostic value of LVSI across all cases and within the four molecular subgroups. This is probably because only presence, and not extent of LVSI was registered in PORTEC-3, and only substantial LVSI has shown to be a strong prognostic factor.^4^

ER status within NSMP EC may also be predictive for response to adjuvant treatment. In this study, we found a small non-significant benefit of CTRT in both ER-positive and ER-negative NSMP EC. Radiotherapy combined with hormonal therapy instead of chemotherapy may be an equally effective but much less toxic alternative for women with high-risk ER-positive NSMP EC. Historical trials did not show a significant benefit of adjuvant hormonal therapy.^41^ However, these trials were done in unselected cohorts, and testing of hormonal therapy specifically among ER-positive NSMP tumours might be the way forward. This will be investigated in the RAINBO NSMP-ORANGE randomized clinical trial (NCT05255653), including women with ER-positive NSMP EC.

In this study, *CTNNB1* exon 3 mutations were not independently associated with recurrence. Previous studies showing an association between *CTNNB1* mutations and adverse clinical outcomes included more women with low- and (high-)intermediate risk EC, potentially explaining the difference in prognostic relevance.^19,20,42^ Also, L1CAM was not an independent predictor for recurrence in our study. Overexpression of L1CAM was most prevalent in the clinically unfavourable p53abn molecular subgroup and did not further deteriorate clinical outcomes in this group. Also within NSMP EC, overexpression of L1CAM was not an independent predictor due to its’ association with negative ER and PR expression. It has been shown that expression of L1CAM is dependent on TGF-signalling and Wnt/-catenin activity, which in turn are inhibited by progesterone.^43-45^

Although we find a strong and independent prognostic impact of ER status in NSMP EC in our study, these findings were not validated in an external validation cohort. However, internal validation using the leave-one-out method and bootstrap resampling confirmed the independent prognostic relevance of ER in NSMP EC. We have investigated the molecular landscape of NSMP EC using IHC and a large targeted NGS panel which showed significant differences between ER-positive and ER-negative NSMP tumours. Investigation of copy number alterations in these tumours could have improved our study as it likely adds molecular and potentially prognostic information.

In conclusion, the prognostic impact of the molecular classification, age, stage, and adjuvant CTRT was confirmed in a large cohort of high-risk EC. The prognostic relevance of tumour grading was limited to NSMP high-risk EC. PR and L1CAM expression and *CTNNB1* mutations had no independent significant prognostic impact. ER-positivity was independently associated with a lower risk of recurrence in NSMP EC and identified a large homogeneous subgroup of NSMP tumours that may represent a novel fifth molecular subgroup. Assessment of ER status in high-risk NSMP EC is feasible in clinical practice and has the potential to improve risk stratification and treatment of patients with NSMP EC.

## Data Availability

Requests for data sharing with a research proposal should be addressed to the corresponding author within 15 years from the date of publication. Depending on the specific research proposal, the TransPORTEC consortium will determine when, for how long, for which specific purposes, and under which conditions the requested data can be made available, subject to ethical consent. 

## Additional Information

## Acknowledgements

We thank all clinical and pathology teams at participating sites of the PORTEC-3, as well as the women who participated in the trials and their families. We thank PORTEC-3 central data manager Karen Verhoeven at the Comprehensive Cancer Center the Netherlands (IKNL) and the international TransPORTEC research consortium. We thank Tessa Rutten, Natalja ter Haar and Enno Dreef (Department of Pathology, Leiden University Medical Center, Leiden, Netherlands) for their technical support. Part of the results of this study was presented at the European Society of Gynaecological Oncology (ESGO) 2021 Congress (October 23-25, 2021, Prague, Czech Republic)^46^ and the United States and Canadian Academy of Pathology (USCAP) 2022 Annual Meeting (March 19-24, 2022, Los Angeles, United States)^47^.

## Author’s contributions

LV, TB, NH, VTHBMS, CLC conceptualized the study. LV, ALC and TB carried out experiments. LV, and NH analysed the data. All authors were involved in writing the paper and had final approval of the submitted and published version.

## Ethics approval and consent to participate

The PORTEC-3 randomized clinical trial was approved by the ethics committees at all participating centres. Written informed consent was obtained from all patients. Translational study of the MST cohort was approved by the Leiden-Den Haag-Delft medical ethics committee, and a waiver for informed consent for the MST cohort was given. The study was performed in accordance with the Declaration of Helsinki.

## Supplementary Information

A Data Supplement (PDF) is available including a detailed description of immunohistochemistry staining and scoring procedures, as well as a detailed description of DNA isolation and sequencing methods. In addition, the Data Supplement contains supplementary figures and tables.

