## Supplementary material for "Prognostic refinement of NSMP high-risk endometrial cancers using oestrogen receptor immunohistochemistry": Data Supplement

### Table of contents

#### Supplementary material and methods

Immunohistochemistry staining procedures…………………………………………....3

Immunohistochemistry staining scoring…………………………………………………...3

DNA isolation and sequencing………………………………………………………………4

#### Supplementary figures

Supplementary Figure S1. Flowchart of patient selection……………………………….5

Supplementary Figure S2. Cohort-specific recurrence-free survival by molecular subgroup……………………………………………………………………………..……....6

Supplementary Figure S3. Recurrence-free survival by histologic subtype and FIGO grade in patients with MMRd, p53abn and NSMP EC……………………………………7

Supplementary Figure S4. Recurrence-free survival of NSMP EC patients by % of ER protein expression..……………………………………………………………………….8

Supplementary Figure S5. Recurrence-free survival of NSMP EC patients by ER and received adjuvant treatment……………………………………………………………...9

Supplementary Figure S6. Incorporation of ER status in the molecular classification of endometrial cancer…………………………………………………………………10

#### Supplementary tables

Supplementary Table S1. Clinicopathological characteristics, by inclusion or not in the translational study…………………………………………………………………........11

Supplementary Table S2. Univariable analysis of recurrence-free survival for MMRd, NSMP and p53abn EC…………………………………………………………..………12

Supplementary Table S3. Internal validation of prognostic value of ER in NSMP EC patients……………………………………………………………………………………13

Supplementary Table S4. Multivariable analysis of recurrence-free survival including a five-class molecular classifier……………………………………...……………...……14

**References** 15

### Supplementary material and methods

#### Immunohistochemistry staining procedures

For each case, one representative formalin-fixed paraffin-embedded tumour block was selected by a pathologist during central pathology review. Immunohistochemistry was performed on 4 µm whole slides. Slides were deparaffinized and rehydrated via graded ethanol series, followed by endogenic peroxidase activity blocking (0.3% Methanol/$H_2O_2$) and antigen retrieval using a microwave oven procedure in 10 mmol/L Tris-EDTA buffer, pH9.0 for 10 minutes. Tissue sections were incubated overnight with primary antibodies against p53 (clone DO-7, 1:2000, DAKO), MLH1 (clone ES05, 1:100, DAKO), MSH2 (clone FE11, 1:100, DAKO), MSH6 (clone EPR3945, 1:800, GENE TEX), ER (clone EP1, 1:400, DAKO), PR (clone PgR636, 1:200, DAKO), and L1CAM (clone 14.10, 1:800, BioLegend) at room temperature or with primary antibody PMS2 (clone EP51, 1:50, DAKO) at 4 degrees. A linker (mouse linker, SM804, DAKO; rabbit linker, SM805, DAKO) was used afterwards for MLH1, PMS2, MSH2 and MSH6. A 30 minute incubation with a secondary antibody (Poly-HRP-GAM/R/R; DPV0110HRP; ImmunoLogic) was then performed. DAB+ (K3468, DAKO) was used as chromogen and sections were counterstained with haematoxylin.

#### Immunohistochemistry staining scoring

For PORTEC-3 cases, MLH1, PMS2, MSH6 and MSH2 protein expression was evaluated to determine MMR status. Tumours were considered MMR deficient if more than 10% of the tumoral nuclei were negative, in the presence of a positive internal control, in at least one of the MMR proteins. For MST cases, immunohistochemistry staining of MMR proteins was performed in a two-stepped approach. Cases with more than 10% loss of PMS2 and/or MSH6 expression were considered MMR proficient. For cases with retained expression of PMS2 and MSH6, additional MLH1 and MSH2 immunohistochemistry was performed to determine final MMR status. Immunohistochemistry for p53 was considered abnormal if more than 10% of the tumour showed strong positive staining of tumour nuclei (overexpression), complete absence of staining with a positive internal control (null-mutant), or significant cytoplasmic staining (cytoplasmic)[1,2]. Immunohistochemistry for ER was considered positive if more than 10% of the tumour showed positive nuclear staining. The cut-off was chosen as it is most commonly used in the assessment of ER expression in endometrial cancer[3-6]. Finally, immunohistochemistry for PR and L1CAM were considered positive when more than 10% of the tumour showed positive nuclear staining. The same as for ER, these cut-off were chosen as they are most commonly used in EC[3,4,7,8]. All immunohistochemistry slides were independently scored by at least two observers (TB, AL, LV). Discrepant results were resolved at simultaneous viewing.

**DNA isolation and sequencing**

Tumour DNA was enriched by taking three 0.8 mm tumoral tissue cores or by microdissection using 5-10 (10) slides on selected tumoral areas by a pathologist, obtaining a tumour percentage >70%. DNA isolation was performed automated using the Tissue Preparation System (Siemens Healthcare Diagnostics). After isolation, the DNA concentration was measured using a fluorometer (Qubit dsDNA HS, Life Technologies, Carlsbad, California, USA). DNA samples were sequenced using the AmpliSeq Cancer Hotspot Panel (Thermo Fisher Scientific, Waltham, MA) version 5 (PORTEC-3) and version 6 (MST). These panels are designed to detect somatic cancer hotspot mutations covering 82 genes, including the complete *POLE* exonuclease domain. Libraries were prepared using 42-84ng of DNA and each sample labelled with a unique barcode (IonCode, TheremoFisher). Ion 540 chips were prepared using Ion Chef System and sequenced using the Ion S5 sequencing System. Sequencing results were evaluated blinded for patient outcome. A minimum coverage threshold of 100 reads and a minimum variant allele frequency of 0.1 reads were considered.

**Supplementary Figure S1. Flowchart of patient selection.**

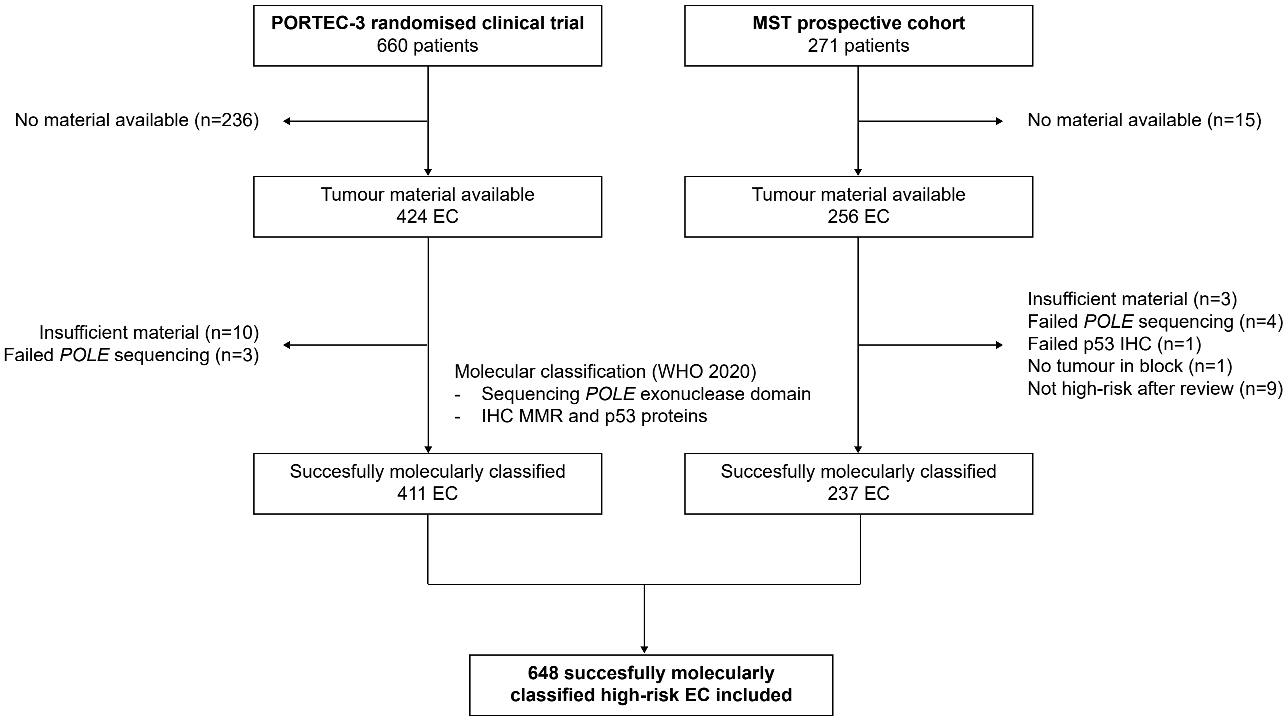


Abbreviations: EC, endometrial cancer; IHC, immunohistochemistry; WHO, World Health Organization; MMR, mismatch repair; HREC, high-risk endometrial cancer.

**Supplementary Figure S2. Cohort-specific recurrence-free survival by molecular subgroup.** Kaplan-Meier survival curves of patients with high-risk endometrial cancer from (A) PORTEC-3 and (B) MST for recurrence-free survival by molecular subgroup.

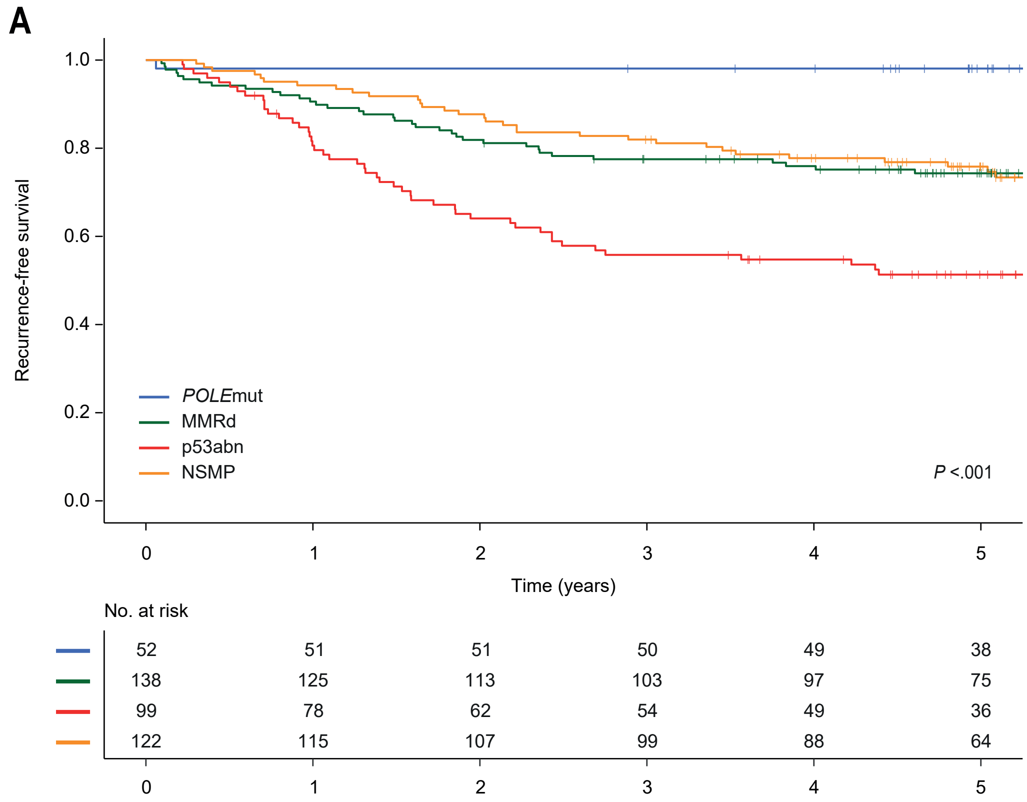


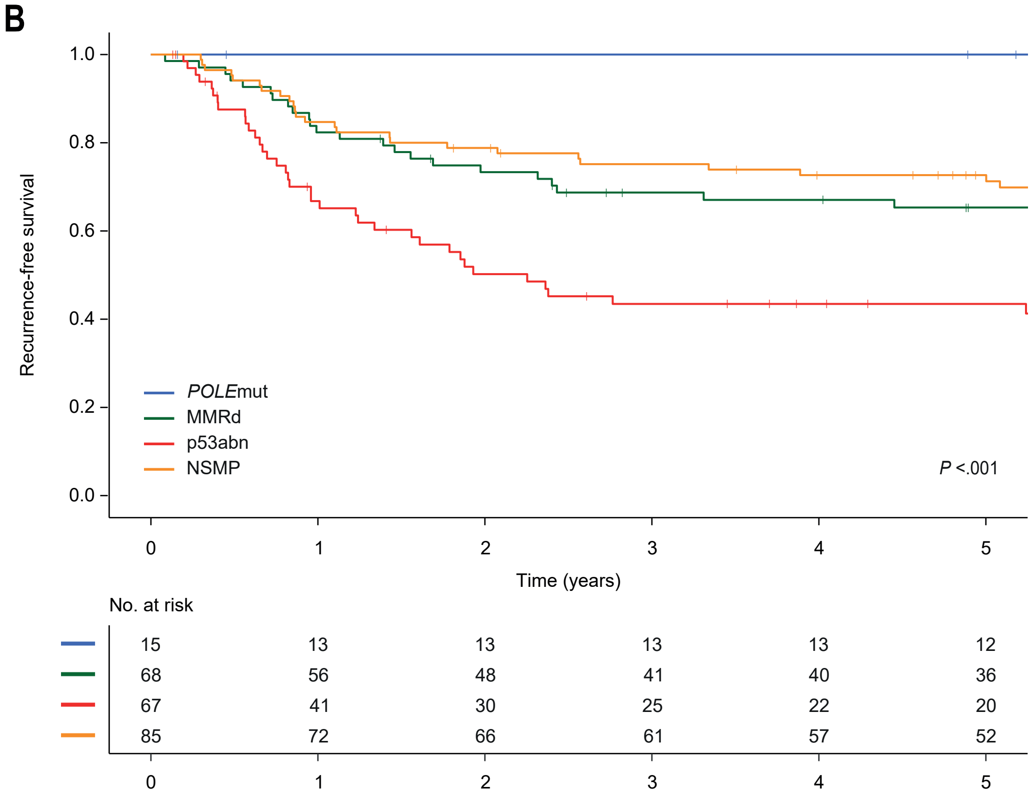

**Supplementary Figure S3. Recurrence-free survival by histologic subtype and FIGO grade.** Kaplan-Meier survival curves of (A) mismatch repair-deficient (MMRd), (B) p53-abnormal (p53abn) and (C) no specific molecular profile (NSMP) high-risk endometrioid endometrial cancer (EEC) for recurrence-free survival by histologic subtype and FIGO grade.

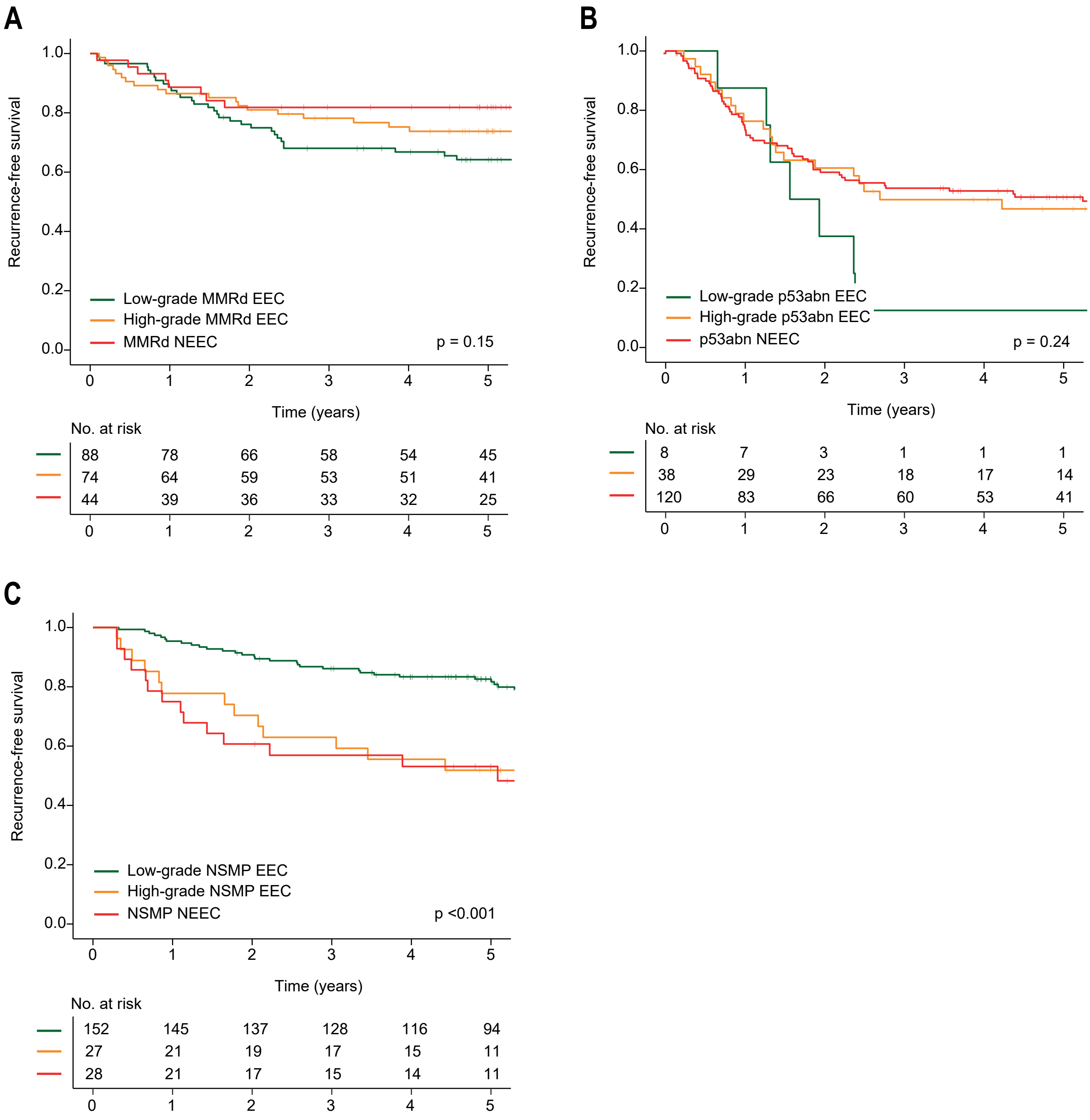

**Supplementary Figure S4. Recurrence-free survival by % of ER protein expression.** Kaplan-Meier survival curves of patients with NSMP high-risk endometrial cancers for recurrence-free survival by different levels of tumour ER protein expression.

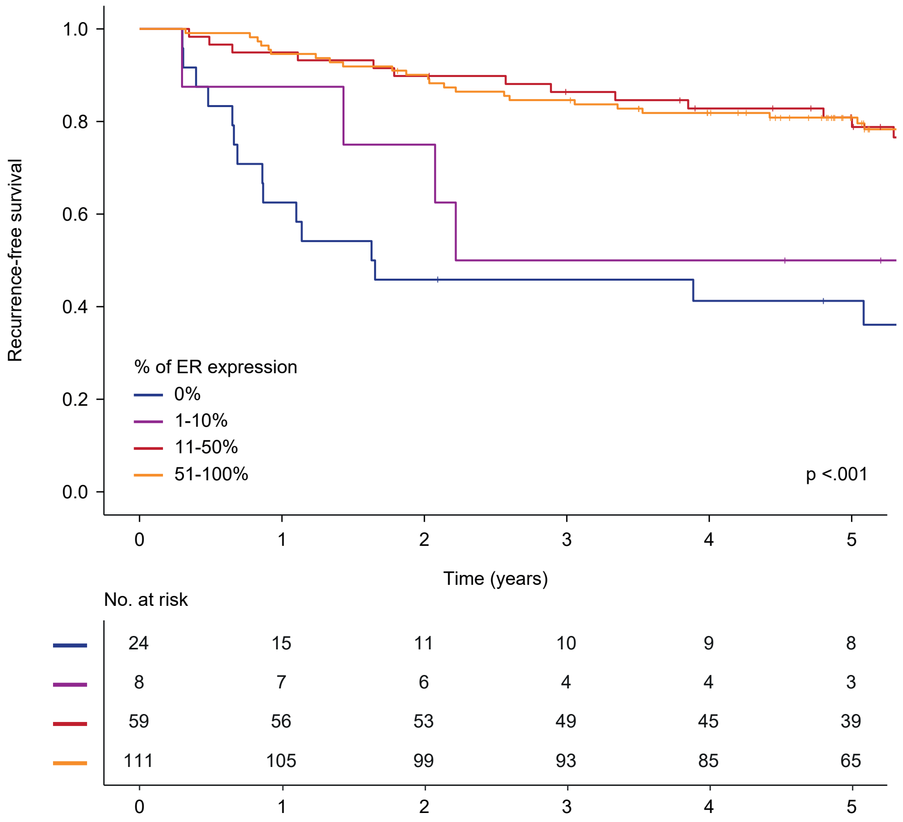

**Supplementary Figure S5. Recurrence-free survival of NSMP endometrial cancer patients by ER status and received adjuvant treatment.** Kaplan-Meier survival curves of patients with NSMP high-risk endometrial cancer for recurrence-free survival by ER status and received adjuvant treatment.

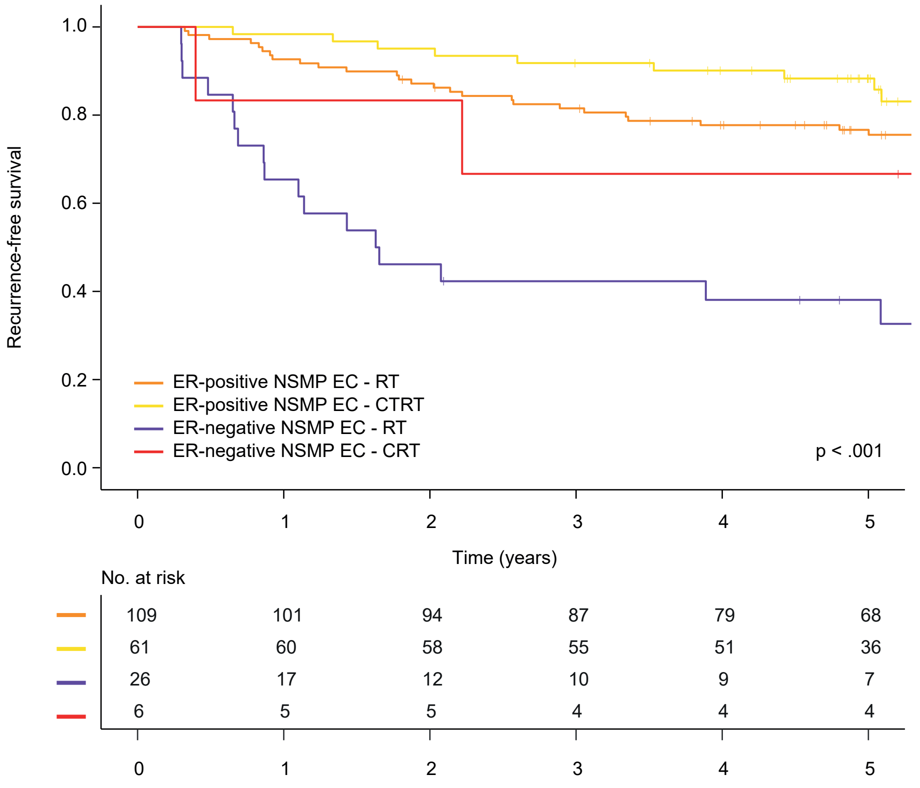

**Supplementary Figure S6. Incorporation of ER status in the molecular classification of endometrial cancer. (A)** Addition of a fourth step into the WHO diagnostic algorithm of the endometrial cancer (EC) molecular classification, including ER immunohistochemistry on NSMP EC. (B) Recurrence-free survival Kaplan-Meier curves of patients with high-risk endometrial cancer.

A

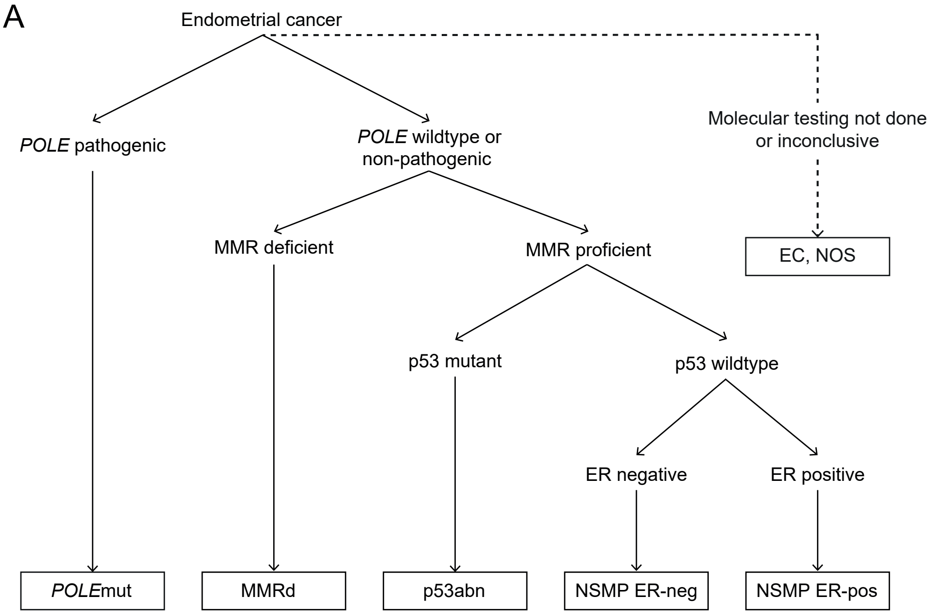


B

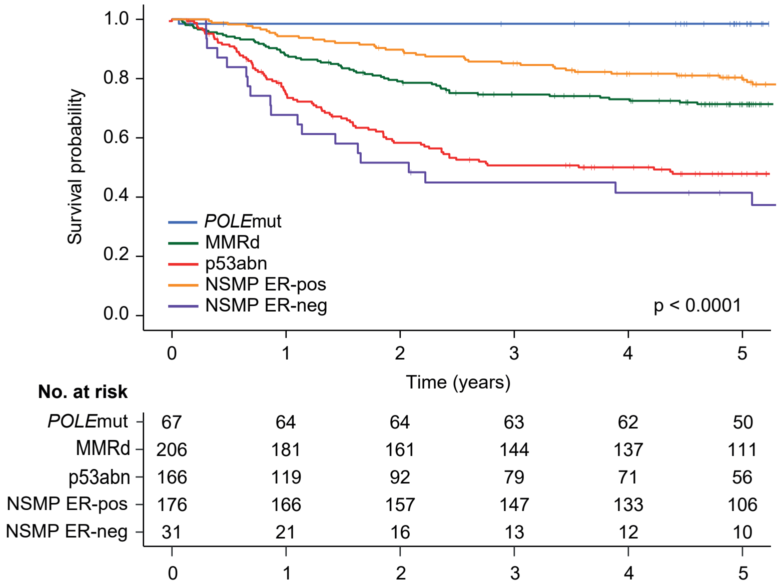


Abbreviations: *POLE*mut, *POLE*-ultramutated; MMRd, mismatch repair deficient; p53abn, p53-abnormal; NSMP, no specific molecular profile; ER-neg, ER-negative; ER-pos, ER-positive; NOS, not otherwise specified.

**Supplementary Table S1. Clinicopathological characteristics, by inclusion or not in the translational study.**

| | **Included** n = 648 (100.0%) | **Excluded** n = 257 (100.0%) | **Total** n = 905 (100.0%) | *p*-value |
|---|---|---|---|---|
| Age | | | | 0.16 |
| Mean (range) | 63.8 (25.0-92.0) | 62.8 (36.1-62.8) | 63.6 (25.0-92.0) | |
| Histotype and grade | | | | 0.27 |
| Low-grade endometrioid | 254 (39.2) | 96 (37.4) | 350 (38.7) | |
| High-grade endometrioid | 179 (27.6) | 74 (28.8) | 253 (28.0) | |
| Serous | 88 (13.6) | 42 (16.3) | 130 (14.4) | |
| Clear cell | 53 (8.2) | 22 (8.6) | 75 (8.3) | |
| Mixed | 31 (4.8) | 14 (5.4) | 45 (5.0) | |
| Carcinosarcoma | 24 (3.7) | 1 (0.4) | 25 (2.8) | |
| Un-/dedifferentiated | 16 (2.5) | 7 (2.7) | 23 (2.5) | |
| Other | 3 (0.5) | 1 (0.4) | 4 (0.4) | |
| Stage | | | | 0.11 |
| IA | 76 (11.7) | 26 (10.1) | 102 (11.3) | |
| IB | 131 (20.2) | 45 (17.5) | 176 (19.4) | |
| II | 181 (27.9) | 68 (26.5) | 249 (27.5) | |
| III | 260 (40.1) | 118 (45.9) | 378 (41.8) | |
| LVSI | | | | 0.16 |
| Absent | 341 (52.6) | 122 (47.5) | 463 (51.2) | |
| Present | 307 (47.4) | 135 (52.5) | 442 (48.8) | |
| Received treatment | | | | 0.001 |
| EBRT | 403 (62.5) | 133 (51.8) | 536 (59.4) | |
| EBRT + CT* | 223 (34.6) | 121 (47.1) | 344 (38.1) | |
| VBT | 19 (2.9) | 3 (1.2) | 22 (2.4) | |
| Follow-up time (years) | | | | <.001 |
| Median (95% CI) | 7.0 (6.7-7.2) | 6.1 (5.9-6.4) | 6.6 (6.3-6.9) | |
| Overall survival | | | | 0.031 |
| 5-year estimate | 71.7% | 77.0% | 73.2% | |

* Including two patients who received VBT+CT.

Abbreviations: LVSI, lymphovascular space invasion; EBRT, external beam radiotherapy; CT, chemotherapy; VBT, vaginal brachytherapy.

**Supplementary Table S2. Univariable analysis of overall recurrence-free survival for MMRd, NSMP and p53abn EC.**

| | | **MMRd EC** | | | | **NSMP EC** | | | | **p53abn EC** | | | |
|---|---|---|---|---|---|---|---|---|---|---|---|---|---|
| | | Total n | HR | 95% CI | *p*-value | Total n | HR | 95% CI | *p*-value | Total n | HR | 95% CI | *p*-value |
| Age | ≤ 60 years | 83 | 1 | | | 85 | 1 | | | 20 | 1 | | |
| | > 60 years | 123 | 1.590 | 0.903-2.798 | 0.11 | 122 | 1.488 | 0.871-2.542 | 0.15 | 146 | 1.103 | 0.565-2.152 | 0.77 |
| Histology and grade | EEC, low-grade | 88 | 1 | | | 152 | 1 | | | 8 | 1 | | |
| | EEC, high-grade | 74 | 0.705 | 0.398-1.248 | 0.23 | 27 | 2.592 | 1.369-4.908 | 0.003 | 38 | 0.573 | 0.243-1.355 | 0.21 |
| | NEEC | 44 | 0.507 | 0.232-1.107 | 0.09 | 28 | 2.939 | 1.575-5.484 | 0.001 | 120 | 0.538 | 0.244-1.187 | 0.13 |
| Stage | I | 60 | 1 | | | 25 | 1 | | | 81 | 1 | | |
| | II | 63 | 1.799 | 0.800-4.049 | 0.16 | 75 | 0.374 | 0.165-0.846 | 0.018 | 32 | 1.821 | 0.980-3.384 | 0.06 |
| | III | 83 | 3.295 | 1.559-6.964 | 0.002 | 107 | 0.827 | 0.412-1.661 | 0.59 | 53 | 4.077 | 2.470-6.732 | <.001 |
| LVSI | Absent | 97 | 1 | | | 123 | 1 | | | 86 | 1 | | |
| | Present | 109 | 1.229 | 0.695-2.174 | 0.48 | 84 | 1.928 | 1.105-3.365 | 0.021 | 80 | 1.448 | 0.927-2.264 | 0.10 |
| Treatment received | EBRT | 136 | 1 | | | 137 | 1 | | | 105 | 1 | | |
| | EBRT+CT | 69 | 1.116 | 0.601-2.072 | 0.73 | 70 | 0.563 | 0.296-1.070 | 0.08 | 59 | 0.675 | 0.402-1.133 | 0.14 |
| ER IHC | Negative (<10%) | 41 | 1 | | | 32 | 1 | | | 81 | 1 | | |
| | Positive (≥10%) | 150 | 1.343 | 0.656-2.748 | 0.42 | 170 | 0.274 | 0.157-0.478 | <.001 | 82 | 1.195 | 0.777-1.838 | 0.42 |
| PR IHC | Negative (<10%) | 79 | 1 | | | 53 | 1 | | | 115 | 1 | | |
| | Positive (≥10%) | 124 | 1.854 | 1.026-3.352 | 0.041 | 149 | 0.395 | 0.234-0.668 | 0.001 | 47 | 0.940 | 0.585-1.510 | 0.80 |
| L1CAM IHC | Negative (<10%) | 176 | 1 | | | 182 | 1 | | | 50 | 1 | | |
| | Positive (≥10%) | 28 | 0.582 | 0.232-1.462 | 0.25 | 24 | 1.856 | 0.929-3.709 | 0.08 | 114 | 1.241 | 0.764-2.016 | 0.38 |
| *CTNNB1* exon 3 | No mutation | 153 | 1 | | | 118 | 1 | | | 144 | 1 | | |
| | Mutation | 19 | 1.018 | 0.402-2.579 | 0.97 | 51 | 0.807 | 0.435-1.497 | 0.50 | 0 | NA | | |

Abbreviations: MMRd, mismatch repair-deficient; EC, endometrial cancer; NSMP, no specific molecular profile; p53abn, p53-abnormal; HR, hazard ratio; CI, confidence interval; EEC, endometrioid endometrial cancer; NEEC, non-endometrioid endometrial cancer; LVSI, lymphovascular space invasion; EBRT, external beam radiotherapy; CT, chemotherapy.

**Supplementary Table S3. Internal validation of prognostic value of ER in NSMP EC patients.**

| | PORTEC-3<br>RFS (32 events) | | | | MST<br>RFS (28 events) | | | |
|---|---|---|---|---|---|---|---|---|
| | Total n | HR | 95% CI | *p*-value | Total n | HR | 95% CI | *p*-value |
| Histology and grade | | | | | | | | |
| Endometrioid, low-grade | 90 | 1 | | | 58 | 1 | | |
| Endometrioid, high-grade | 13 | 2.10 | 0.78-5.70 | 0.14 | 13 | 2.54 | 0.86-7.50 | 0.09 |
| Non-endometrioid | 14 | 1.58 | 0.41-6.08 | 0.50 | 14 | 1.44 | 0.37-5.6 | 0.60 |
| Stage | | | | | | | | |
| I-II | 52 | 1 | | | 45 | 1 | | |
| III | 65 | 2.04 | 0.95-4.36 | 0.07 | 40 | 2.31 | 1.01-5.31 | 0.048 |
| Treatment received* | | | | | | | | |
| RT (VBT or EBRT) | 55 | 1 | | | NP | | | |
| RT (VBT or EBRT) + CT | 62 | 0.50 | 0.24-1.02 | 0.06 | | | | |
| ER IHC | | | | | | | | |
| Negative (<10%) | 13 | 1 | | | 19 | 1 | | |
| Positive (≥10%) | 104 | 0.31 | 0.09-1.06 | 0.06 | 66 | 0.32 | 0.10-1.03 | 0.06 |

* Analysis of MST patient not corrected for treatment as the majority of patients (93.1%) were treated with radiotherapy alone.

Abbreviations: RFS, recurrence-free survival; HR, hazard ratio; CI, confidence interval; RT, radiotherapy; VBT, vaginal brachytherapy; EBRT, external beam radiotherapy; CT, chemotherapy; IHC, immunohistochemistry; NP, not performed.

**Supplementary Table S4. Multivariable analysis of overall recurrence-free survival including a five-class molecular classifier.**

| **Recurrence** | **Multivariable analysis** | | |
|---|---|---|---|
| **n = 643, 207 events** | **HR** | **95% CI** | ***p*-value** |
| Age | | | |
| ≤60 years | 1 | | |
| >60 years | 1.39 | 0.99-1.95 | 0.06 |
| Stage | | | |
| I | 1 | | |
| II | 1.54 | 0.99-2.41 | 0.06 |
| III | 3.25 | 2.21-4.76 | <.001 |
| Histology and grade | | | |
| Endometrioid, low-grade | 1 | | |
| Endometrioid, high-grade | 1.15 | 0.75-1.76 | 0.52 |
| Non-endometrioid | 1.01 | 0.64-1.60 | 0.97 |
| LVSI | | | |
| Absent | 1 | | |
| Present | 1.29 | 0.95-1.76 | 0.10 |
| Treatment received | | | |
| RT (VBT or EBRT) | 1 | | |
| RT (VBT or EBRT) + CT | 0.68 | 0.48-0.95 | 0.023 |
| Molecular subgroups | | | |
| MMRd | 1 | | |
| *POLE*mut | 0.12 | 0.03-0.50 | 0.003 |
| p53abn | 2.82 | 1.90-4.19 | <.001 |
| ER-positive NSMP | 0.69 | 0.45-1.06 | 0.09 |
| ER-negative NSMP | 2.27 | 1.33-3.90 | 0.003 |

Model fit multivariable model: Akaike's information criterion (AIC) 2162.38, model concordance (C-index) 0.726.

Abbreviations: HR, hazard ratio; CI, confidence interval; LVSI, lymphovascular space invasion; RT, radiotherapy; VBT, vaginal brachytherapy; EBRT, external beam radiotherapy; CT, chemotherapy; MMRd, mismatch repair-deficient; *POLE*mut, *POLE* ultra-mutated; p53abn, p53-abnormal; NSMP, no specific molecular profile.
